# Automatic segmentation and quantification of nigrosome-1 neuromelanin and iron in MRI: a candidate biomarker for Parkinson’s disease

**DOI:** 10.1101/2023.04.13.23288519

**Authors:** Mikel Ariz, Martín Martínez, Ignacio Alvarez, Maria A. Fernández-Seara, The Catalonian Neuroimaging Parkinson’s disease Consortium, Pau Pastor, Maria A. Pastor, Carlos Ortiz de Solórzano

## Abstract

The dopaminergic neuronal loss in the substantia nigra pars compacta (SNc) has been related to a reduction of neuromelanin (NM) and accumulation of iron in the nigrosome-1 (N1) in Parkinson’s disease (PD). This suggests that N1 degeneration could be a promising early biomarker of PD. To date, only qualitative visual scales have been used to assess its degeneration in iron-sensitive images. Here we present the first fully-automatic method for the quantification of NM and iron content in the N1. Our method uses a multi-image atlas populated with healthy N1 structures that implements a customised label fusion strategy to segment the N1. NM-MRI and susceptibility-weighted images (SWI) of 71 PD patients and 30 healthy controls (HCs) were used in the study. Our quantification showed that N1’s NM content was reduced and the iron content increased in PD patients compared with HCs. ROC analyses showed the high diagnostic potential of N1, and revealed that the N1 alone was more sensitive than the entire SNc to detect abnormal iron accumulations in PD patients. Multi-parametric binary logistic regression showed that computer-assisted diagnosis methods could benefit from the segmentation of the N1 to boost their performance. A significant correlation was also found between most N1 image parameters and both disease duration and the motor status scored with the Unified Parkinson’s disease rating scale part III (UDPRS-III), suggesting a NM reduction along with an iron accumulation in N1 as the disease progresses. In addition, voxel-wise analyses revealed that this association was stronger for the N1 than for the entire SNc, highlighting the benefits of an accurate segmentation of the N1 to monitor disease course.

## 1. INTRODUCTION

Parkinson’s disease (PD) is caused by the loss of dopaminergic neurons in the substantia nigra pars compacta (SNc). Its incidence increases with age, and it is one of the major causes of disability. PD early diagnosis is still a challenge as it is based on the clinical assessment of the subject discarding other potential causes of parkinsonism, and the patient’s positive response to Levodopa therapy, leading to a misdiagnosis rate of approximately 16% (Adler et al., 2014).

The substantia nigra (SN), located in the ventral tegmentum of the midbrain, is divided into two main regions: the iron-rich ventral SN pars reticulata (SNr), and the dorsal SNc, where the dopaminergic neurons containing neuromelanin (NM) are located. NM is believed to have a neuroprotective function against the toxicity of iron-mediated oxidative processes (Zecca et al., 2008; Zucca et al., 2017), and the death of dopaminergic neurons of the SNc causes a NM depigmentation, followed by an increase of iron load (Sulzer et al., 2018). Indeed, we and others have previously observed a decrease of SNc NM in PD patients compared with healthy controls (HC) (Castellanos et al., 2015; Sasaki et al., 2006). In a more recent study, we simultaneously observed a decrease of SNc NM along with an increase of SNc iron in PD patients compared with HCs (Martínez et al., 2023), in line with previous findings by others (He et al., 2021; Langley et al., 2020; Takahashi et al., 2018).

The SNc can be divided, based on calbindin immunostaining, into matrix and five clusters of high NM concentration, labelled as nigrosomes 1-5 (Damier et al., 1999a). The nigrosome-1 (N1), located in the dorsolateral region of the SNc, is the largest of the nigrosomes and the one that is affected first and the most in PD, with up to 98% of its NM-rich dopaminergic neurons lost (Damier et al., 1999b). N1 is usually visualized using T2* weighted imaging or susceptibility weighted imaging (SWI) (Cheng et al., 2020; Schwarz et al., 2014). In images of healthy adult brains, the N1 appears as a hyperintense structure in the dorsolateral region of the SN. The *N1 sign* is thus visible as a bifurcated hypointense tail with a hyperintense middle, also called the “swallow-tail” sign (Kim et al., 2019).

The fact that N1 is suffering the loss of dopaminergic neurons in PD long before any clinical symptoms are developed makes it a promising potential biomarker for early PD diagnosis (Damier et al., 1999b). The disappearance of the N1 sign in PD has been associated with the loss of dopaminergic NM-rich neurons and increased iron deposition in the lateral-ventral region of the SNc (Blazejewska et al., 2013; He et al., 2022; Huddleston et al., 2017; Jin et al., 2019; Langley et al., 2017; Schwarz et al., 2018). Recent studies have focused specifically on assessing the degeneration of N1 in PD (Langley et al., 2020), and evaluating the potential of the N1 sign as a diagnostic biomarker for PD (Chau et al., 2020; Cheng et al., 2020; He et al., 2021; Jokar et al., 2023; Mahlknecht et al., 2017; Pavese & Tai, 2018). However, all previous works have either indirectly measured the N1 degeneration by describing NM-reduced areas that only partially overlap with the N1 (Langley et al., 2020), or have evaluated the diagnostic performance of the N1 sign through a binary, qualitative visual assessment of “no loss”, “unilateral loss” or “bilateral loss” of the structure (Cheng et al., 2020; He et al., 2021; Jokar et al., 2023). Therefore, the anatomic segmentation of the N1 would allow to accurately quantify the NM reduction and increased iron deposition associated with PD, and to further develop a robust, automatic diagnostic tool for PD.

In this study, we aim to fill this gap by proposing the first fully-automatic method for the segmentation of N1 in SWI images that allows an accurate localization of this region inside the SNc. Since the method is based on a multi-image atlas, it detects the N1 region in a robust manner, even if the N1 sign has been bilaterally lost due to PD progression. We use our automatic segmentation to quantify N1 NM and iron in HCs and PD patients to measure the N1 degeneration in PD. Furthermore, we assess the diagnostic potential of N1 as a PD biomarker in comparison with the entire SNc, and we investigate the specific association of N1 image features with clinical variables of the disease.

## 2. METHODS

### 2.1. Subjects

HC subjects and PD patients belonged to a cohort used in a previous work (Martínez et al., 2023). The cohort was composed of 71 PD patients and 30 age and sex-matched HCs, recruited by the Catalonian Neuroimaging Parkinson’s disease Consortium, who joined the study between April 2018 and October 2020. This study was approved by the University of Navarra Research Ethics Committee, and written informed consent was obtained from all subjects. PD patients were clinically diagnosed according to the UK Brain Bank Parkinson’s disease criteria (Hughes et al., 1992). Patients underwent an interview covering demographic data, family history of neurological diseases, the Mini-Mental State Examination (MMSE), the Unified Parkinson’s Disease Rating Scale Part-III (UPDRS-III) and the Modified Hoehn & Yahr Scale (H&Y). Disease onset was determined at the age at which the Parkinsonian motor signs started, either self-reported or reported by the caregiver. The UPDRS-III was assessed in the ON state right before the MRI scan.

### 2.2. MRI data acquisition

MRI scans were collected on a 3T MAGNETOM Skyra MRI scanner (Siemens Healthineers, Erlangen, Germany) using a 32-channel head coil. An anatomical T1-weighted image, a NM-MRI sequence and a SWI sequence were collected in a 32-minute session. The NM-MRI images were obtained with a 3D-NM sensitive T1-weighted turbo spin-echo sequence (Nakane et al., 2008), and the SWI images were obtained combining a long-TE high-resolution fully flow-compensated 3D GRE sequence with filtered phase information in each voxel (Haacke et al., 2009). Four scans of the NM sequence and three scans of the SWI sequence were acquired and averaged offline to account for head motion artifacts. Further acquisition details can be found in our previous study (Martínez et al., 2023). The final voxel sizes of the NM-sensitive images and SWI images were 0.57×0.57×1 mm^3^ and 0.69×0.69×2 mm^3^, respectively. To facilitate and standardize manual delineations, all images were manually reoriented based on the axial and mid-sagittal plane of a canonical T1 template image. All preprocessing was done using SPM12 (The Wellcome Centre for Human Neuroimaging, UCL Queen Square Institute of Neurology, London, UK) and custom scripts in Matlab (The MathWorks, Inc., MA, USA).

### 2.3. Segmentation of nigrosome-1

The image processing workflow for the automatic segmentation of N1 was developed based on a 3D atlas-based pipeline proposed in our previous works (Ariz et al., 2019; Martínez et al., 2023). In our current work, we maintain an atlas-based registration scheme, in which the target image is individually registered with a set of images that compose the atlas and have an associated label of N1. We created one atlas of NM images and one atlas of SWI images, composed of the NM and SWI images of the HCs, respectively, and their associated label images. The N1 was manually delineated in the SWI images by an experienced neurologist (MA Pastor). The criterion to annotate the N1 was to find an ovoid hyperintense region in the dorso-lateral aspect of the caudal SNc and below the red nucleus. The SNc was also manually delineated by the same expert in NM-MRI images.

The registration of the target image with each of the atlas images was carried out in three steps: first, an affine transformation was calculated for the whole brain as an initial, coarse alignment. Second, a volume of 130×140×46 voxels and 110×120×26 voxels that contain the brainstem in the NM and SWI image, respectively, was automatically cropped, and a subsequent affine registration that accounts for global brainstem size and shape differences was carried out. And third, an elastic B-spline transformation was iteratively optimized, which accounts for local size and shape variations. All registrations were performed using a multiresolution strategy with three resolution levels to accelerate computation, and mutual information was maximized (Mattes et al., 2003) using an adaptive stochastic gradient descent optimizer (Klein et al., 2009). All transformations were calculated with Elastix, an open-source software for image registration (Klein et al., 2010).

The calculated transformations were then sequentially applied to the corresponding atlas labels in a label propagation stage to obtain as many segmentation candidates as images composing the atlas. The final segmentation step was the fusion of the labels, in which a final 3D segmentation mask was obtained from all the candidate labels. We compared three different label fusion strategies: majority voting (MV), global weighted voting (GWV), and local weighted voting (LWV). The three of them were implemented in Matlab.

All segmentations were evaluated by means of the dice similarity coefficient (DSC). To assess the accuracy of N1 segmentation –and of the SNc-in an objective manner, a leave-one-out strategy was adopted with the pool of HCs, in which each of the 30 controls was segmented using an atlas composed of the remaining 29 HCs, and the DSCs between the automatic segmentation masks and the manual delineations were calculated. Then, the 71 PD patients were also segmented using the NM and SWI atlases of 30 HCs.

#### 2.3.1. Label fusion based on Majority Voting

MV is by far the most typical label fusion strategy and the one we adopted in our previous works (Ariz et al., 2019; Martínez et al., 2023). In MV, all the candidates have the same weight in the decision, thus each voxel in the final segmentation mask is calculated as the mode of the distribution of values of the corresponding voxel throughout all the candidate mask images. The voxel will be labelled correctly if the majority of the candidates agree on the correct label.

#### 2.3.2. Label fusion based on Global Weighted Voting

GWV is a label fusion strategy in which each candidate has a different weight in the voting, based on the similarity between the image being segmented –i.e., the target image- and the corresponding member of the atlas. The weight is global in the sense that it is unique for all the voxels of the candidate. MV can be seen as a specific case of GWV, in which all the candidates have equal, unitary weights. We propose a specific adaptation of GWV to our problem with the aim of taking advantage of *a priori* knowledge. First, MV is applied to automatically segment the entire brainstem in the target image. We then use this segmentation to calculate a similarity measure between the target image and each of the atlas images constrained to the brainstem region, instead of using the whole image volume. This strategy avoids possible image artifacts outside the brainstem affecting the similarity calculation, such as aliasing or ghosting, and constrains the similarity measure to the actual region of interest for PD.

There are different similarity metrics that can be used to obtain a measure of how similar two images –or in this case, image regions-are, such as the mean square distance, variants of the correlation coefficient, or the mutual information (Mattes et al., 2003). Different assumptions are made when each of them is used for image registration (Roche et al., 2000). In this case, we chose to use the normalized mutual information (NMI), which is the one that best accounts for the intensity variations that exist between MR images of different subjects, and the NM and iron structure heterogeneity in PD. The weight *w* that we assign to each candidate in GWV is thus defined as:

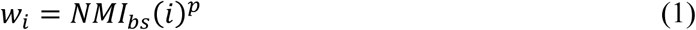

where *NMI* _*bs*_(*i*) is the NMI between the brainstem region of the atlas image *i* and the target image, and *p* is a gain exponent that allows to maximize weight differences between segmentation candidates when the NMI is not sensitive enough. We empirically set *p* to 10 and 12 for the NM-MRI and SWI sequence, respectively. Following this definition, the more similar the brainstem region between the target and the atlas image is, the higher the weight of that atlas element will be in the voting process of candidate labels to obtain the final segmentation mask of the target image.

#### 2.3.3. Label fusion based on Local Weighted Voting

The last label fusion strategy we implemented is LWV, in which each voxel can receive a different weight value based on a local similarity measure. Here again, *a priori* knowledge is used to optimize the process. MV is calculated to segment the entire brainstem in the target image and the application of LWV is constrained to the voxels contained in the brainstem. A local neighbourhood is defined around each brainstem voxel, in which a similarity measure between the target image and each atlas image is calculated and used to obtain the weight of the voxel. The local weight of each voxel *v* in LWV is calculated as:

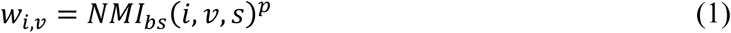

where *NMI* _*bs*_(*i,v,s*) is the NMI between the atlas image *i* and the target image for a window of *s*×*s* pixels around the voxel *v* contained in the brainstem, and *p* is a gain exponent that maximizes weight differences between segmentation candidates when the NMI is not sensitive enough. We maintained the values *p*= 10 and *p*= 12 chosen for GWV, and we empirically set *s* to 31 and 21 pixels for the NM-MRI and SWI sequence, respectively.

### 2.4. NM and SWI image registration

To measure both NM and iron content in N1, an intra-subject NM and SWI sequence alignment was first carried out. To this end, a multimodal image registration pipeline was designed: a rigid transformation was calculated between each subject’s NM and iron-sensitive images through a multiresolution registration with four resolution levels, using an adaptive stochastic gradient descent optimizer (Klein et al., 2009) to maximize the mutual information (Mattes et al., 2003). The intra-subject alignment accuracy was assessed by measuring the DSC of the manual delineations of the brainstem between the NM image and the transformed SWI image.

### 2.5. Quantification of NM and iron in nigrosome-1

The automatic segmentation of N1 in SWI images and the intra-subject alignment of NM and SWI sequences allowed us to quantify the amount of NM and iron contained in the N1 both in HC and in PD. Two image quantification measures were calculated for NM and iron: the contrast ratio (CR) and the normalized volume (nVol). The CR was measured as the percentage increase of the N1 mean intensity with respect to the brainstem mean intensity, and can thus be interpreted as the normalized brightness of N1. The nVol was calculated as the volume of N1 divided by the total grey matter volume to account for brain size differences among subjects. Since iron appears as hypo-intense in SWI images, iron CR values were inverted in sign to facilitate the reader’s interpretation, meaning that higher iron CR values correspond to darker regions in the image.

Following what was done in our previous work, we thresholded both NM and iron images, and measured CR and nVol only for hyper-intense (NM) or hypo-intense (iron) voxels contained in the N1 segmentation mask. The optimal thresholds for N1 (i.e., the ratio by which a N1 voxel’s intensity exceeds, in the case of NM, or is lower than, in the case of iron, the average brainstem intensity) were found after sweeping a range of threshold values with the aim of maximizing HC vs. PD discrimination, choosing the threshold for which the p-value of a Mann-Whitney’s U test between HC and PD was minimum (see **Supplementary Fig. 1**). The chosen threshold values were 0.14 and 0.22 for NM CR and NM nVol, respectively, and 0 and 0.20 for iron CR and iron nVol, respectively.

### 2.6. Creation of NM and iron probabilistic maps for nigrosome-1

All the images were registered to a reference coordinate space, specifically to the space defined by the subject with the highest DSC for N1 segmentation. All SWI images were first aligned with their corresponding NM images following the procedure described in Section 2.4, and then all the images (both NM and SWI) were transformed to the reference space following the three-step multiresolution strategy described in Section 2.3. The automatic N1 segmentation masks underwent the same sequence of transformations. The aligned N1 masks were used to create a N1 template mask in the reference space, and spatial probabilistic maps of NM and iron content in the N1 in HC and PD were created by assigning to each N1 voxel the number of subjects that contain –according to our optimal thresholds-NM or iron in that voxel. The probabilistic maps were finally normalized with respect to the number of subjects in each group (N=30 in HC and N=71 in PD).

### 2.7. Statistical analysis and diagnostic performance

As most of the distributions of the measured parameters did not meet the normality assumption, nonparametric tests were used for comparisons. Specifically, a Mann-Whitney’s U test was used to assess the statistical significance of age differences between HCs and PD patients, and a chi-square test for sex differences. The three segmentation methods proposed were compared by means of a Friedman test and post hoc analysis with Bonferroni correction. NM and iron quantification values between HCs and PD patients were compared using a Mann-Whitney’s U test. Receiver operating characteristic (ROC) analysis was used to evaluate the diagnostic performance of the quantified NM and iron parameters through their corresponding area under the curve (AUC) values. Different parameters were combined in single –optimal-models by means of binary logistic regression, and these models were also evaluated using their associated ROC curves. Partial correlation analysis was used to find significant relationships between each quantified image parameter and the clinical variables (i.e., disease duration and UPDRS-III) in the PD group. The Spearman’s rank correlation coefficient (i.e., ρ) was calculated for each image-clinical parameter combination. Age and sex were used as covariates in the analysis, since they both affect the phenotypical expression of PD (Prange et al., 2019), and p-values were adjusted for comparisons through a multivariate permutation test (Groppe, 2022). Voxel-wise Spearman’s partial correlation coefficients were also calculated between the NM and iron CR of each SNc voxel and the clinical scores, using also sex and age as covariates and the multivariate permutation test to adjust the p-values to account for multiple voxels and multiple clinical scores, to investigate the spatial patterns of the correlation between the image signal and the clinical variables. The distribution of the correlation coefficients of all N1 voxels was then compared with the one of all SNc voxels for each NM/iron – clinical score combination by means of a Mann-Whitney’s U test. All tests were two-tailed, and corrected *p* values <0.05 were considered statistically significant. All statistical analyses were performed using the Statistics and Machine Learning Toolbox of Matlab.

### 2.8. Data and code availability statement

The data and code reported in this study will be made publicly available upon acceptance of the manuscript. In the meantime, they will be available for reviewers upon request to the corresponding author C. Ortiz de Solórzano.

## 3. RESULTS

Demographic and clinical characteristics of the subjects are described in Table 1. There were no statistically significant differences in sex or age between HC and PD groups.

Mean ± standard deviation and [range] values are represented in the table. The UPDRS-III was measured while PD patients were in the ON medicated state. Statistical significance of HC and PD differences was measured by means of a Mann-Whitney’s U test for age and a chi-square test for sex.

**Table 1.** Demographic and clinical characteristics of HC and PD groups

|  | Healthy controls | PD patients | p-value |
| --- | --- | --- | --- |
| **Age** | 62.73 ± 7.83 [51-81] | 65.34 ± 10.32 [39-83] | 0.11 |
| **Sex (F/M)** | 17/13 | 34/37 | 0.42 |
| **Disease duration (years)** | n/a | 8.86 ± 5.39 [2-23] | - |
| **UPDRS-III** | n/a | 20.57 ± 11.63 [2-61] | - |
| **Hoehn and Yahr scale** | n/a | 2.07 ± 0.54 [1-5] | - |
| **Most affected side (L/R/B)** | n/a | 39/30/2 | - |

### 3.1. Segmentation of N1

The results of the automatic segmentation of N1 and SNc are summarized in **Fig. 1**. The best segmentation of N1 was achieved using GWV combination strategy, with an average DSC of 0.642 for the 30 SWI images of the HCs. Both GWV and LWV performed significantly better than MV for the task of N1 segmentation. In the case of SNc segmentation, MV performed significantly better than GWV. The DSC of brainstem segmentation in NM images was also calculated as a control, with both MV and GWV performing significantly better than LWV.

**Figure 1.**
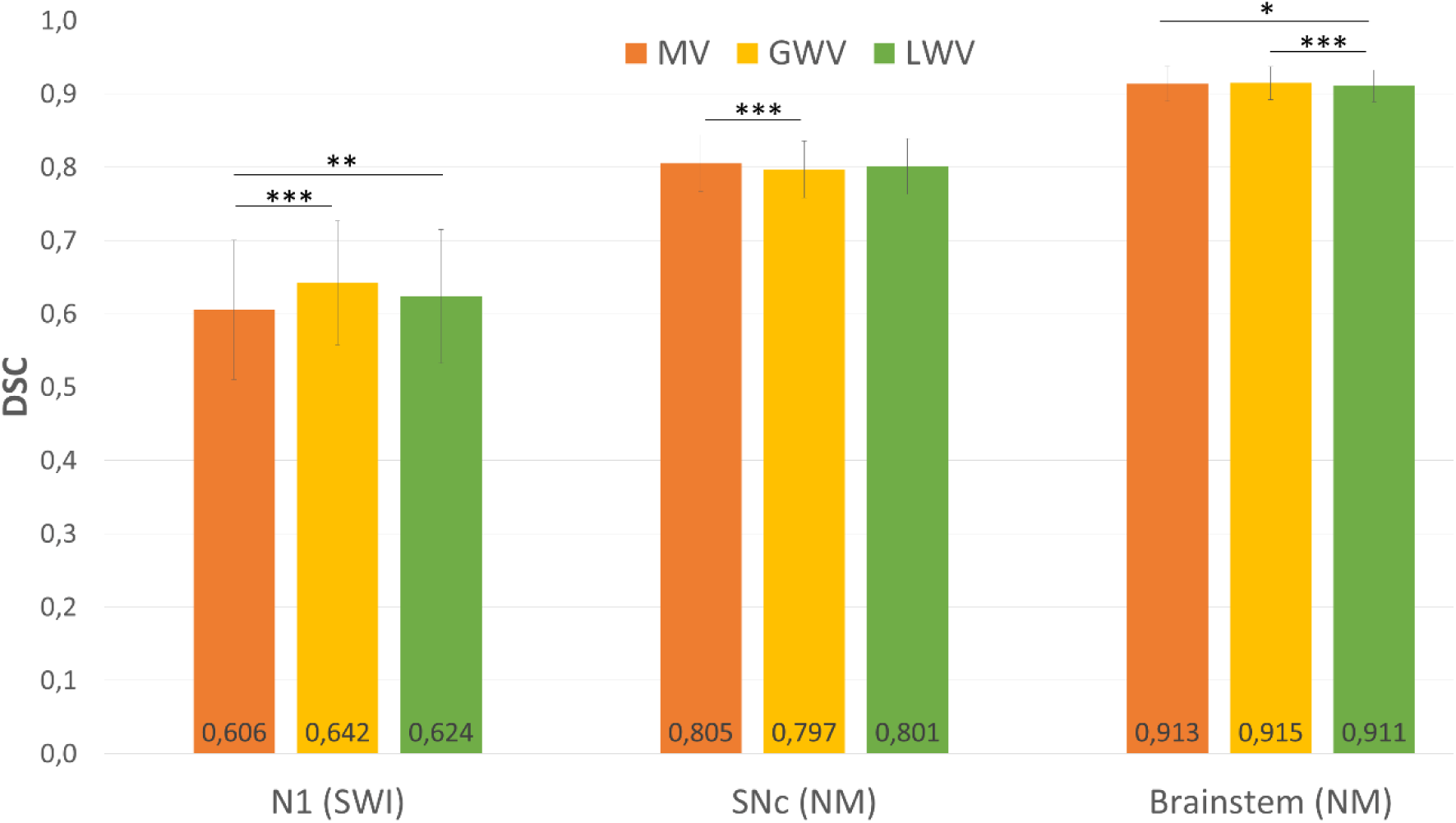
Automatic segmentation results for N1 and SNc. The DSC values obtained by the three label fusion methods proposed (MV, GWV and LWV) on the N1 in SWI images, and the SNc and brainstem in NM-MRI images, are shown. Statistically significant differences between methods for the segmentation of each structure were measured by means of a Friedman test and post hoc analysis with Bonferroni correction (* p<0.05, ** p<0.01, *** p<0.001).

Globally, GWV is the label fusion method that yields the highest segmentation accuracy for the N1 while performing well for the rest of the structures. Using GWV, we achieve DSCs of 0.642, 0.797 and 0.915 for the segmentation of N1, SNc and brainstem, respectively.

Representative segmentation examples of a HC, a PD patient with a well-preserved N1 sign, and a PD patient with the N1 sign completely lost are shown in **Fig. 2**. N1 segmentation contours are shown in cyan for a 2D slice of the SWI image of each subject (**Fig. 2b,f,j**), together with the corresponding raw slice (**Fig. 2a,e,i**) to better appreciate the complexity of the task, especially when the N1 sign is completely lost (**Fig. 2i,j**), as the structure is not visible and a manual delineation of the N1 contour would be unfeasible. The segmented N1 contour is also overlaid on the NM image, previously aligned with the SWI image, and displayed together with the SNc segmentation in magenta (**Fig. 2d,h,l**), and the corresponding raw slice (**Fig. 2c,g,k**). Brainstem segmentation contours are also displayed in yellow.

**Figure 2.**
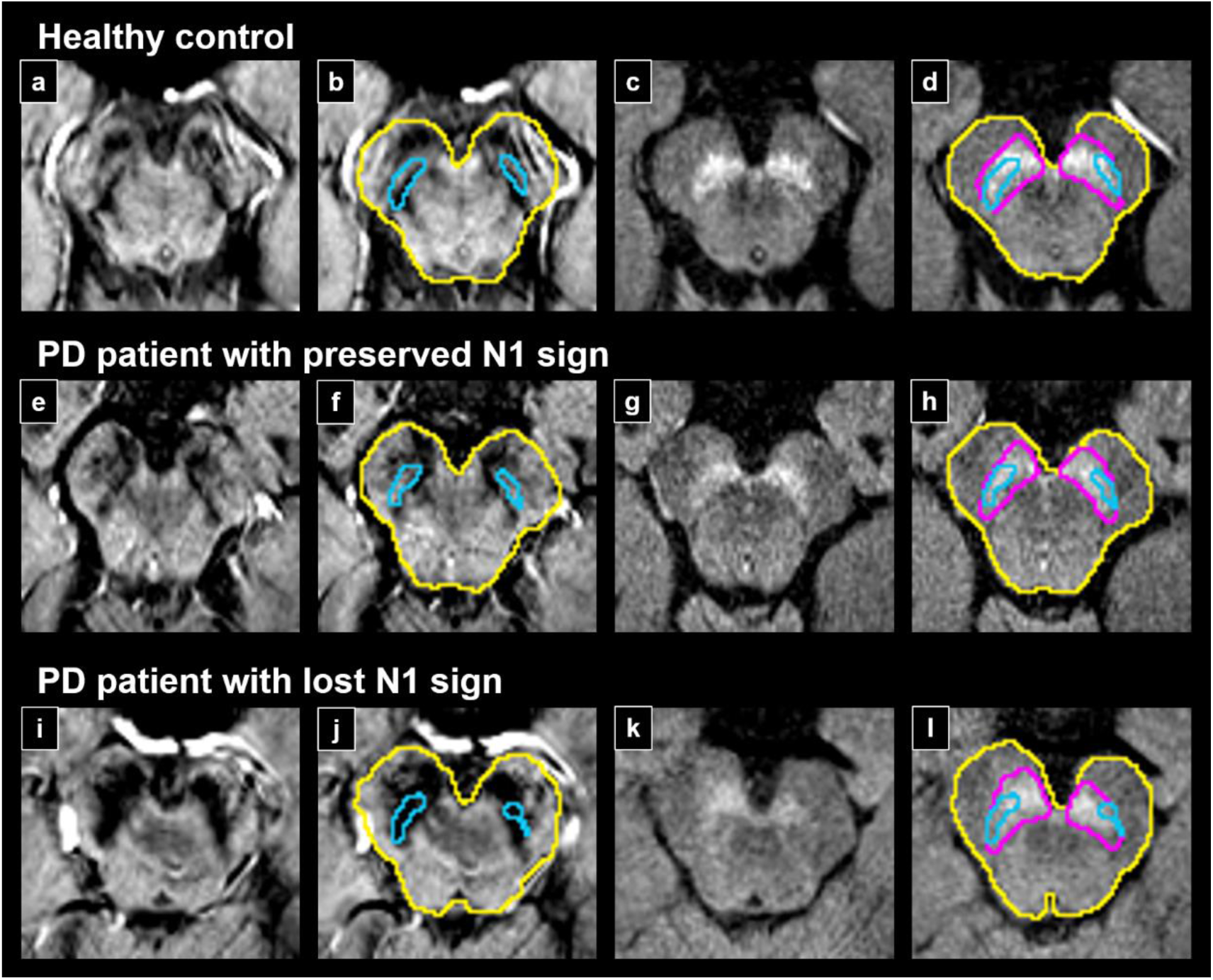
Example segmentations of N1 and SNc. Representative segmentations of a healthy control (a-d), a PD patient with the N1 sign preserved (e-h), and a PD patient with the N1 sign lost (i-l) are shown for NM and SWI images after sequence alignment. Axial 2D slices with segmentations of N1 in cyan (b,f,j) with their corresponding raw SWI slices (a,e,i) are displayed, as well as segmentations of SNc in magenta with the N1 region overlaid in cyan (d,h,l) with their corresponding raw NM slices (c,g,k). Brainstem segmentations are shown in yellow. Note that the N1 sign is completely lost in the bottom-row example and a manual delineation of the structure would be impossible.

### 3.2. NM-MRI and SWI spatial alignment

Each subject’s SWI and NM-MRI images were registered and placed in a common space so that the N1 mask obtained from the SWI images could also be applied to the NM image for NM quantification. The comparison of the brainstem annotations of both images, averaged across all HC and PD subjects, yielded a DSC of 0.927 ± 0.012 as a measure of the intra-subject alignment accuracy.

### 3.3. NM and iron quantification in N1

NM and iron quantification results for N1 and SNc are summarized in **Fig. 3**. PD patients showed a strong reduction of N1 NM CR and N1 NM nVol (p<0.001, purple violins in **Fig. 3**), along with an increase of N1 iron CR and N1 iron nVol (p<0.001, cyan violins in **Fig. 3**) compared with HCs. A similar behaviour was observed for the NM and iron content of the SNc. PD patients showed a reduction of SNc NM CR and SNc NM nVol (p<0.001, magenta violins in **Fig. 3**) along with an increase of SNc iron CR and SNc iron nVol (p<0.001, yellow violins in **Fig. 3**) compared with HCs.

**Figure 3.**
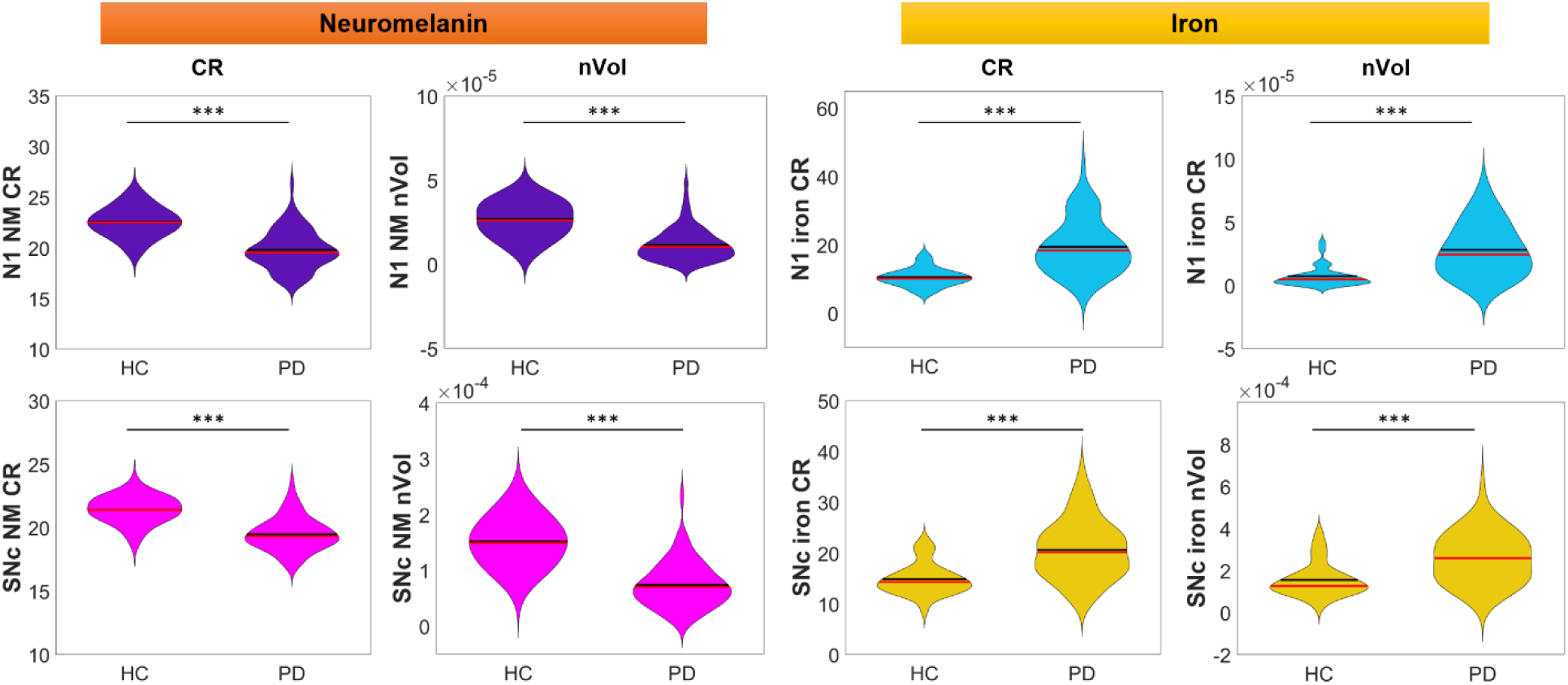
NM and iron quantification results of HC and PD groups for N1 (top row) and SNc (bottom row). CR and nVol measured for N1 NM (top row in purple), N1 iron (top row in cyan), SNc NM (bottom row in magenta) and SNc iron (bottom row in yellow) are shown. Statistically significant differences between groups were measured by means of Mann–Whitney’s U test (* p<0.05, ** p<0.01, *** p<0.001).

Spatial probabilistic maps of NM and iron content in N1 for HC and PD groups are shown in **Fig. 4** The probability of each N1 voxel containing NM and/or iron is visualized using a specific heatmap, and the average brainstem (in dark grey) and SNc (in light grey) are also drawn as spatial references. A higher N1 NM content in HCs with respect to PD patients is visually confirmed by the warmer colours shown in their respective probability maps, which indicates a higher probability of a N1 voxel containing NM if it belongs to a HC rather than to a PD patient. In a similar way, a lower N1 iron content in HCs with respect to PD patients is visually confirmed by the colder colours of the HCs’ iron probability maps, which indicates that a N1 voxel of a PD patient has a higher probability of containing iron than a N1 voxel of a HC.

**Figure 4.**
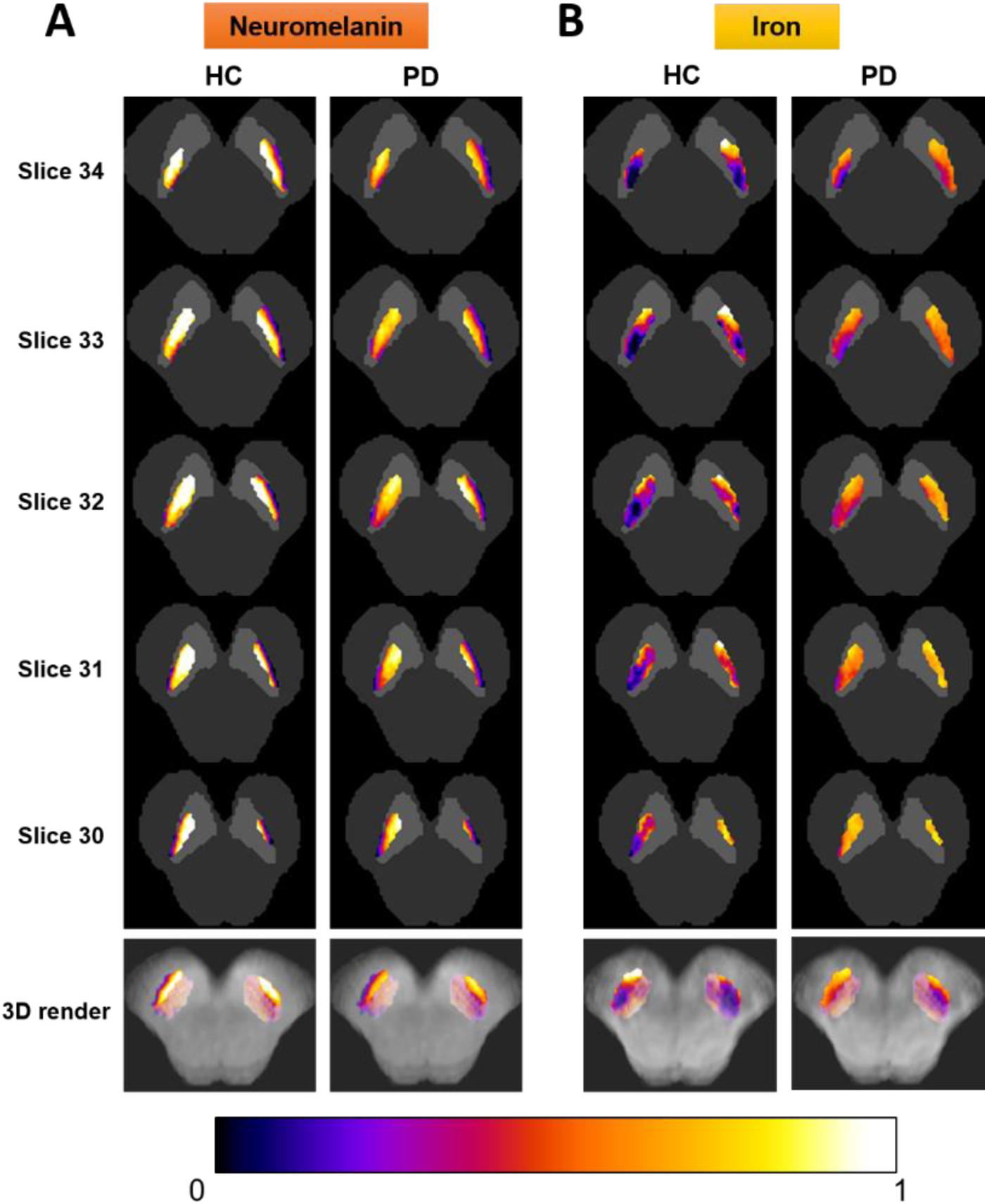
Spatial probabilistic maps of NM and iron content in N1 for HC and PD. 2D axial views of the five slices that contain the average N1 template are shown, with the probability of each N1 voxel containing NM (A) and/or iron (B) represented with a specific heatmap (warm colours for high probabilities and cold colours for low probabilities). Average brainstem (in dark grey) and SNc (in light gray) masks are shown as spatial references. A 3D volumetric render of the N1 spatial probability map is also shown in the bottom row. To improve visual quality, all maps were spatially upsampled by a factor of four using bicubic interpolation.

### 3.4. Diagnostic performance

Diagnostic performance results for the discrimination between HCs and PD patients using the quantified image parameters of the N1 and the SNc are summarized in **Fig. 5** and **Supplementary Table 1**. First, all the quantification parameters were assessed individually by means of their corresponding ROC curves and the associated AUC values (**Fig. 5A**). Then, different quantification parameters were combined through binary logistic regression to assess more complete models (**Fig. 5B**).

**Figure 5.**
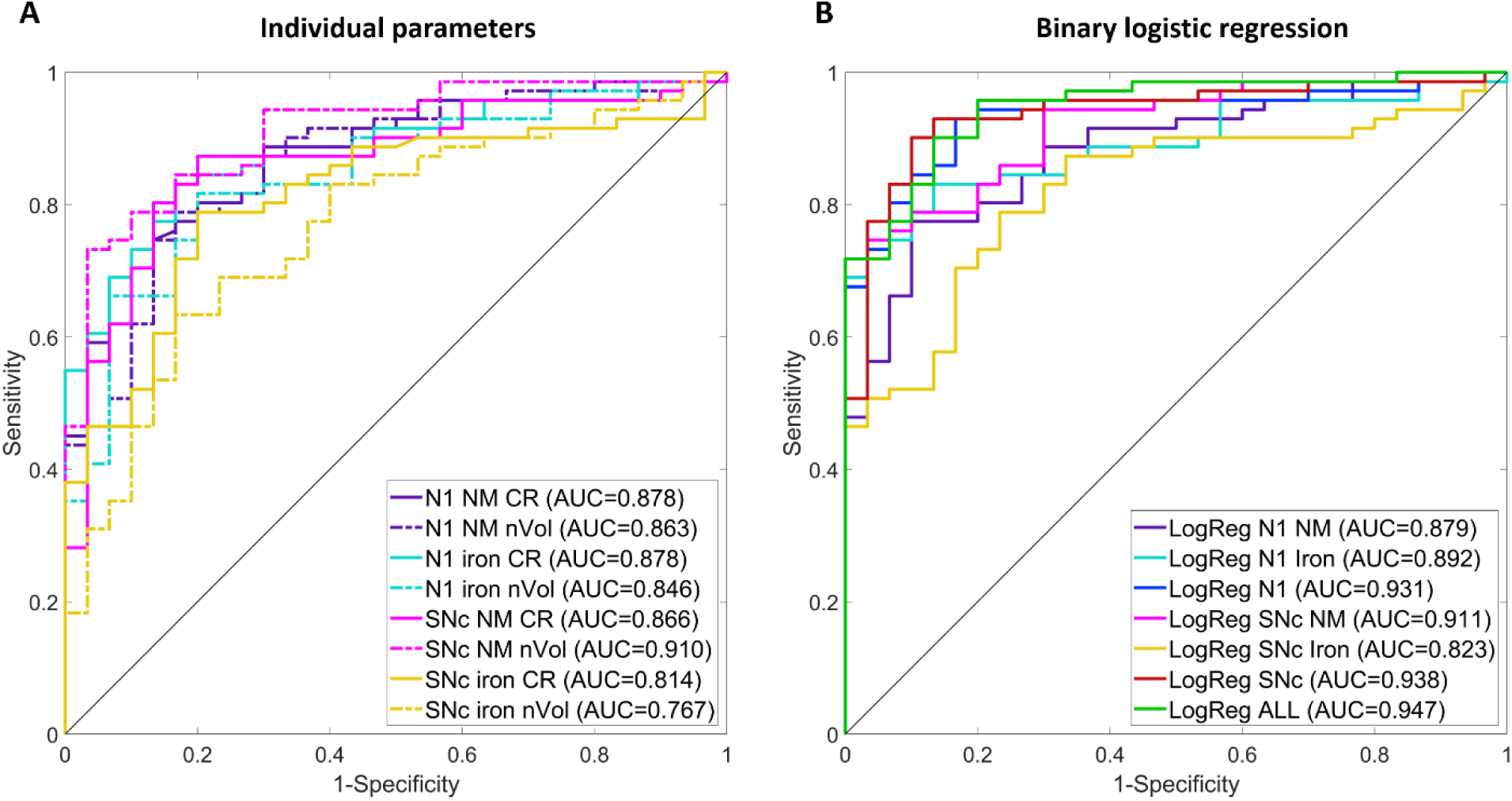
Diagnostic performance results for HC vs. PD discrimination using image parameters automatically calculated for N1 and SNc. A) ROC curves and AUC values of quantified individual image parameters (CR and nVol) of N1 (NM and iron) and SNc (NM and iron). B) ROC curves and AUC values of binary logistic regression models combining different image parameters. The following models were created combining CR and nVol parameters: N1 NM (purple), N1 iron (cyan), complete N1 combining NM and iron (blue), SNc NM (magenta), SNc iron (yellow), complete SNc (red), and a final complete model including all N1 and SNc parameters (green). Abbreviations: LogReg, logistic regression.

The analyses revealed that N1 was the structure with the highest, most consistent discrimination power both for NM and iron, with AUC values of 0.879 and 0.892, respectively, when CR and nVol were combined. The SNc, on the other hand, showed the highest discrimination power for NM, with an AUC of 0.911, but a lower discrimination power for iron, with an AUC of 0.823. When both NM and iron parameters were combined in a single –optimal-model for each structure, N1 and SNc showed very similar ability to distinguish between HC and PD, with AUCs of 0.931 and 0.938, respectively. Finally, when both structures were combined in a final, complete model, the AUC was boosted to 0.947.

### 3.5. Correlation analysis between N1 image features and clinical scores

Partial correlation analyses revealed that most of N1 image quantification parameters were significantly correlated with clinical scores (**Supplementary Fig. 2**). Specifically, N1 NM CR was reduced with disease duration (ρ = -0.31, p = 0.010), whereas an increase in both N1 iron CR (ρ = 0.43, p < 0.001) and N1 iron nVol (ρ = 0.46, p < 0.001) was associated to a longer disease duration. A similar behaviour was observed for UPDRS-III, since N1 NM nVol decreased for larger UPDRS-III values (ρ = -0.28, p = 0.021), whereas both N1 iron CR (ρ = 0.33, p = 0.006) and N1 iron nVol (ρ = 0.25, p = 0.041) increased with UPDRS-III. There was also a statistical trend towards a negative correlation between N1 NM nVol and disease duration (ρ = -0.21, p = 0.089), and between N1 NM CR and UPDRS-III (ρ = -0.24, p = 0.052).

Interestingly, voxel-wise partial correlation analyses revealed a higher association of clinical scores with NM and iron voxels of N1 than with the entire SNc voxels (**Fig. 6**). Indeed, a significantly stronger negative correlation between NM and both disease duration and UPDRS-III was found for N1 voxels than for SNc voxels (**Fig. 6C**), meaning that this relationship is mainly localized in the N1 region of the SNc. In a similar way, a significantly stronger positive correlation between iron and both clinical scores was found for N1 voxels than for SNc voxels (**Fig. 6C**). This behaviour can also be qualitatively observed in the 2D axial slices of the voxel-wise correlation maps between NM and iron CR, and disease duration (**Fig. 6A**) and UPDRS-III (**Fig. 6B**). The Spearman’s partial correlation coefficient between each SNc voxel’s CR and the clinical score is shown by means of a blue/red colormap, where blue indicates a strong negative correlation and red indicates a strong positive correlation. The N1 region inside the SNc is outlined in cyan. It can be visually observed that, in general, stronger correlations are found for N1 voxels than for the rest of SNc voxels. Limiting the visualization to those voxels for which the correlation between the CR and the clinical variable is statistically significant (p<0.05) supports this finding, especially for iron (**Supplementary Fig. 3**).

**Figure 6.**
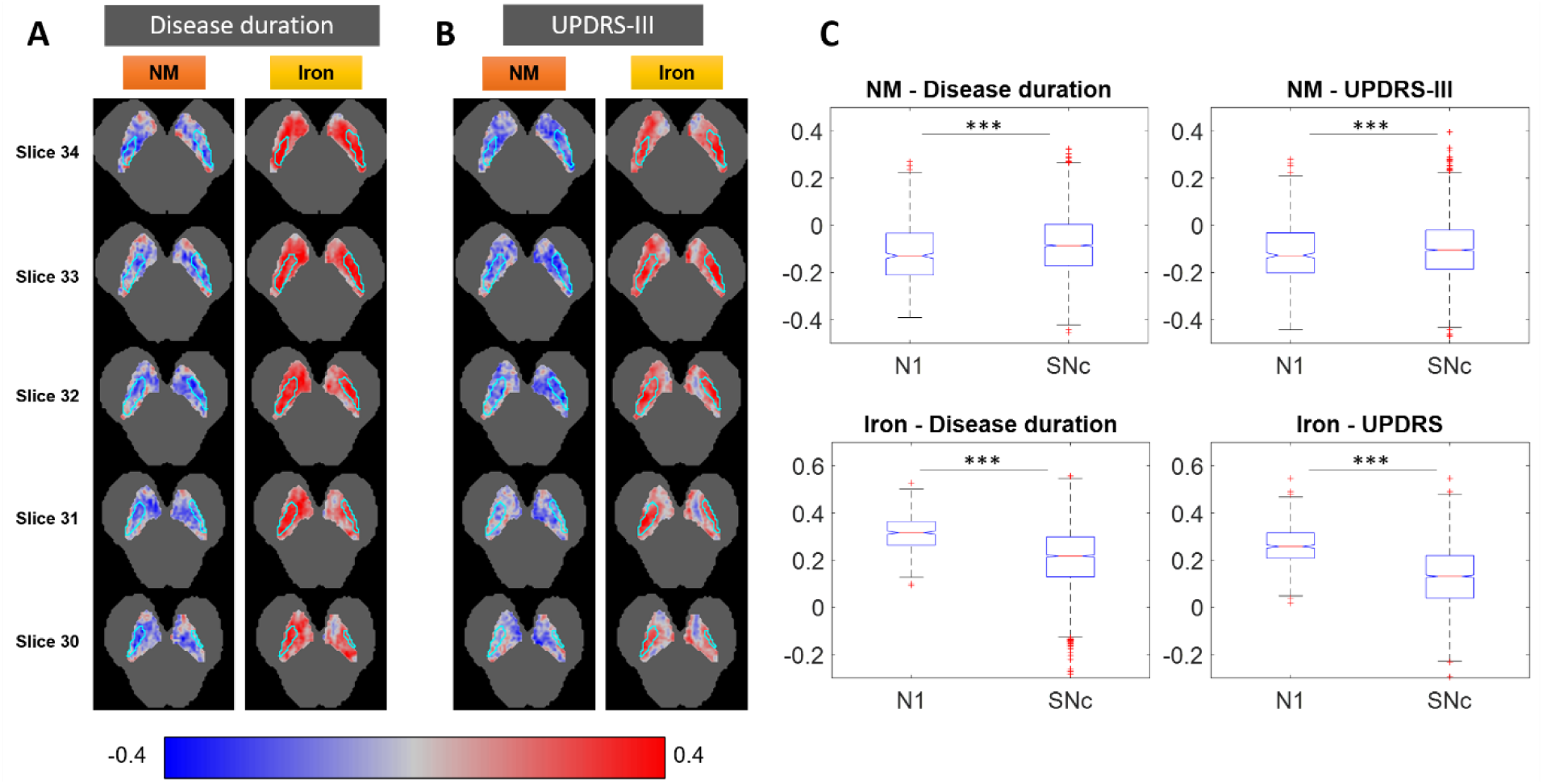
Voxel-wise correlations between clinical scores and NM and iron in the N1 and SNc. 2D axial slices of voxel-wise correlations between the CR of NM and iron of each SNc voxel, and disease duration (A) and UPDRS-III (B). The N1 region is outlined in cyan, and the brainstem template is shown in gray as reference. Partial correlation analysis was performed using age and sex as covariates, and a multivariate permutation test was used to adjust the p-values to account for multiple voxels and multiple clinical scores. A blue/red colormap is used to display the value of the Spearman’s rank partial correlation coefficient of each voxel. Red indicates positive associations between voxel signal and clinical score, whereas blue indicates negative associations. C) Boxplots comparing the distribution of the correlation coefficients of all N1 voxels with the one of all SNc voxels. Statistically significant differences were assessed by means of a Mann-Whitney’s U test (* p<0.05, ** p<0.01, *** p<0.001).

## 4. DISCUSSION

To the best of our knowledge, we have presented the first, fully-automatic method for the segmentation of N1, which is a very promising biomarker for the early diagnosis of PD (Chau et al., 2020; Cheng et al., 2020; He et al., 2021, 2022; Jokar et al., 2023). Indeed, recent studies have shown the potential of the N1 sign as a PD biomarker, as the loss of this sign has been associated with higher iron content in that area (Cheng et al., 2020), and PD patients with the highest iron content usually present bilateral loss of the N1 sign (He et al., 2021). However, the N1 sign *per se* does not quantitatively represent either NM or iron in the N1. Moreover, there is some overlap between PD patients and age-matched HCs in terms of the N1 image appearance – i.e., signal intensity-that the N1 sign, a “black or white” measure, is not able to account for. Our fully automatic N1 segmentation method fills this gap, providing a tool that retrieves the specific anatomical location of N1 in SWI images and opens the door to the accurate quantification of NM and iron in the structure. Since our segmentation method is based on the use of an anatomical atlas –i.e., it uses the annotations of the N1 in an atlas of HCs for the segmentation of a new image through image registration, label propagation and label fusion-, it detects the N1 region in a robust manner, even if the N1 sign has been bilaterally lost due to the progression of the disease. In those cases, even a manual delineation of the N1 by an expert neurologist would be unfeasible.

We implemented three label fusion methods –MV, GWV and LWV- and compared them for the task of N1 segmentation. The methods based on weighted voting (GWV and LWV) seemed *a priori* more appropriate for this segmentation problem, given the reduced size and diffuse signal of the N1 (Artaechevarria et al., 2009). These methods were adapted to calculate the weights based only on brainstem local similarities, eliminating the effect of image artifacts located outside it. The complete segmentation pipeline is fully automatic with no need of user interaction –not even the delineation of an initial region of interest-. Segmentation evaluation showed that GWV is the best option for N1 detection, with a top accuracy (DSC) of 0.642. This is consistent with a previous study (Artaechevarria et al., 2009) that showed that global weighting label fusion is more appropriate than local weighting for the segmentation of structures of diffuse appearance that display relative low contrast with respect to their surrounding tissue, as it is the case of N1 in SWI. Taking into account the small size, diffuse nature and shape variability of N1 between subjects, a DSC∼0.64 obtained with GWV can be considered very sensitive. To our knowledge, this is the first attempt at segmenting the N1, thus no performance comparison can be carried out. Recent studies on SNc segmentation, though, have reported DSCs similar to the ones achieved by us for this structure, i.e., ∼0.8 (Gaurav et al., 2022; Le Berre et al., 2019). Both of these previous studies implemented the U-Net, a convolutional neural network (CNN) that learns image intensity patterns from a set of training images to automatically segment the SNc on new unseen images. However, this type of deep learning strategy would likely fail to detect the N1 on most PD patients, for which the structure is hardly visible and the patterns learnt by the network are no longer apparent in the image. Since our method is based on the projection of healthy structures on the target image, after the alignment of the atlas images with the target following a robust multiresolution registration procedure, it is not affected to the same extent by those significant intensity pattern variations and hence performs robustly, even finding the N1 region when it is invisible to the expert’s eye.

N1 nVol and CR quantification showed a very significant (p<0.001) NM reduction along with an increase of iron content in PD patients in comparison with HCs, which is an analogous behaviour to that observed for the SNc. These results are in accordance with what the literature has already suggested on the N1. Some studies have visually assessed, using an ordinal scale, the NM and iron content in the N1 in PD (Jin et al., 2019; Schwarz et al., 2018), whereas others have measured a NM loss and iron increase in the SNc in PD that was especially pronounced in the lateral-ventral part of the SNc (Huddleston et al., 2017; Langley et al., 2017), probably related with the N1 (Langley et al., 2020). Moreover, He et al. (He et al., 2021) found that the PD subjects with bilateral loss of the N1 sign had the highest iron content in the SNc. Our quantification results are also visually reflected in the NM and iron probabilistic maps of the N1 for the two groups, where the N1 sign loss is apparent for PD, and in the ROC analyses for diagnostic performance. The N1 and SNc individually, combining their NM and iron CR and nVol through binary logistic regression, achieved almost identical AUCs when discriminating HCs from PD patients (AUC∼0.93), but the N1 showed a more consistent discrimination power overall for both NM and iron measures separately –i.e., AUCs of 0.88 and 0.89-, whereas the SNc showed a high discrimination power for NM –i.e., AUC of 0.91-but a moderate discrimination power for iron –i.e., AUC of 0.82-. A combined regression model including the SNc and N1 parameters boosted the diagnostic performance to a top AUC of 0.95. These results suggest that the variation of iron content in the N1 rather than in the entire SNc represents better the affection of PD, and any computer-assisted diagnosis model may benefit from a specific segmentation of the N1 region.

Finally, we found correlation between most of the N1 image parameters and clinical variables of PD, which points at the potential of the N1 to monitor the disease. For most parameters, NM content is negatively associated with both disease duration and UPDRS-III, whereas iron content is positively correlated with them, which suggests a NM reduction along with an iron accumulation in N1 as the disease progresses and the motor symptoms worsen. Interestingly, the voxel-wise correlation analyses revealed that this association between clinical scores and image features is stronger in the N1 than in the entire SNc, which further supports the interest in having a specific, accurate segmentation of the N1 region, and extols the value of the method presented in this paper.

We acknowledge some limitations in our study. Currently, most studies aiming at quantifying iron content through MRI utilize quantitative susceptibility mapping (QSM) rather than SWI (He et al., 2022). QSM models each voxel so that its intensity correlates with a physiological magnitude of iron concentration independent from the acquisition device or settings, and we acknowledge that QSM is the most sensitive technique to small changes in iron concentration (He et al., 2015). Even though using SWI images we cannot obtain absolute quantitative iron concentration values, by normalizing their intensities with respect to the brainstem intensity we are indeed able to compare the same anatomical hypointense regions across subjects between HCs and PD patients, demonstrating a –relative-abnormal iron accumulation in the N1 in PD. Future studies using QSM along with our N1 segmentation algorithm on other PD patient cohorts should be carried out to further confirm our findings and better assess N1’s potential as a PD biomarker. QSM being more sensitive than SWI to small iron concentration changes, N1’s diagnostic performance could even increase. Moreover, N1 being the first SNc area affected by PD, a cohort of early-stage PD patients would specifically assess its potential as a biomarker for early-stage or prodromal diagnosis, since our segmentation tool would provide the means to analyse local, subtle changes in NM and iron concentration in the N1.

In summary, we have presented the first method for the segmentation of N1 in iron-sensitive MRI to quantify specific NM and iron content variations caused by PD. The method is fully automatic and robust, able to detect the N1 region even if the N1 sign is completely lost. Our analyses suggest that the N1 is more sensitive than the entire SNc to iron content accumulations caused by PD, and that any computer-assisted diagnosis method would benefit from a specific segmentation of the N1 to boost its performance. Our method also provides the means to detect local, subtle changes in NM or iron content and assess the real potential of N1 as a biomarker for the early diagnosis of PD in future studies.

## Data Availability

The data and code reported in this study will be made publicly available upon acceptance of the manuscript. In the meantime, they will be available for reviewers upon request to the corresponding author C. Ortiz de Solorzano.

## Abbreviations

AUC: area under the curve
CR: contrast ratio
DSC: dice similarity coefficient
GWV: global weighted voting
HC: healthy control
LWV: local weighted voting
MV: majority voting
N1: nigrosome-1
NM: neuromelanin
NMI: normalized mutual information
nVol: normalized volume
PD: Parkinson’s disease
SN: substantia nigra
SNc: substantia nigra pars compacta
SNr: substantia nigra pars reticulata
SWI: susceptibility-weighted imaging
UPDRS-III: unified Parkinson’s disease rating scale part-III.

## CRediT author contribution statement

**Mikel Ariz:** Conceptualization; Writing – Original draft; Writing – Review & Editing; Methodology; Software; Investigation; Formal analysis; Data curation; Visualization. **Martín Martínez:** Conceptualization; Writing – Review & Editing; Methodology; Investigation; Formal analysis; Data curation; Visualization. **Ignacio Alvarez:** Investigation; Data curation. **María A. Fernández-Seara:** Methodology; Investigation. **The Catalonian Neuroimaging Parkinson’s disease Consortium:** Resources. **Pau Pastor:** Conceptualization; Writing – Review & Editing; Resources; Supervision; Project administration; Funding acquisition. **María A. Pastor:** Conceptualization; Writing – Review & Editing; Data curation; Resources; Supervision; Project administration; Funding acquisition. **Carlos Ortiz de Solórzano:** Conceptualization; Writing – Review & Editing; Methodology; Supervision; Project administration; Funding acquisition.

## Acknowledgements

The authors would like to thank all the subjects who participated in this study. RTI2018-094494-B-C22 and PDI2021-122409OB-C22 (to C.O.S) and SAF2016-81016-R (to M.A.P) grants from the Spanish Ministry of Science and Innovation and Universities (MCIU/AEI/FEDER, UE).

