## Supplementary Material for "Automatic segmentation and quantification of nigrosome-1 neuromelanin and iron in MRI: a candidate biomarker for Parkinson’s disease"

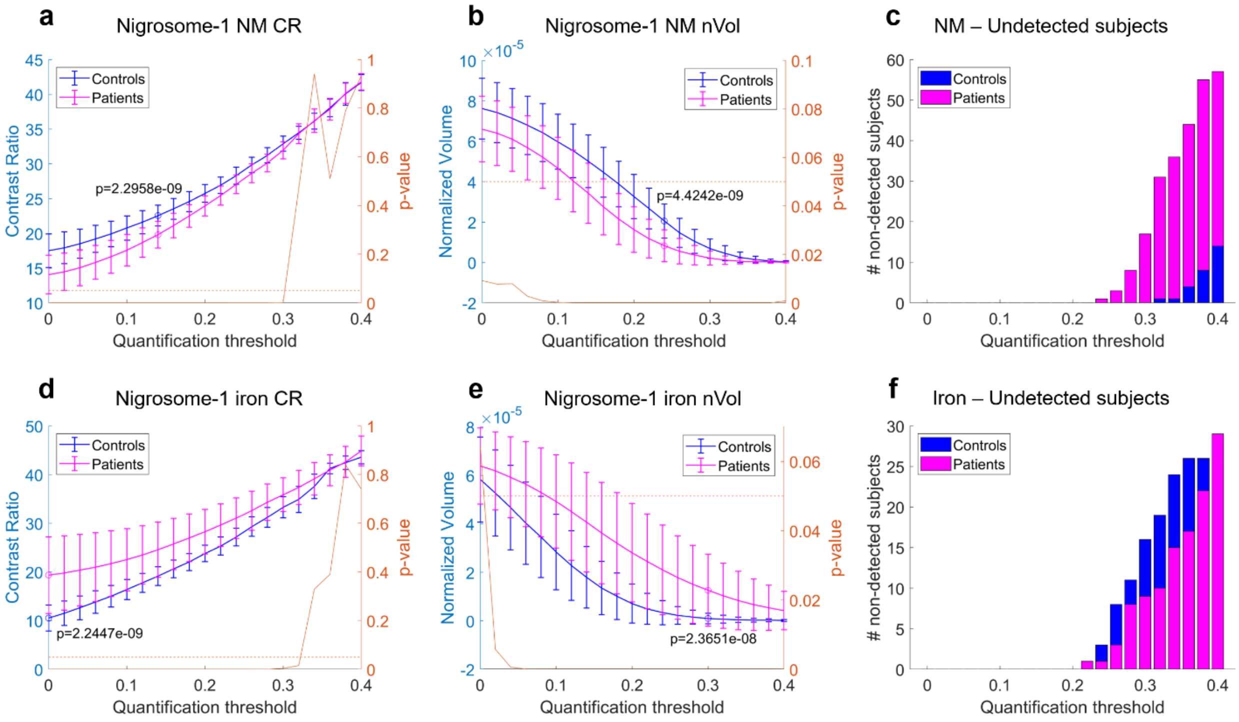


**Supplementary Figure 1. Quantification threshold analysis.** Analysis of neuromelanin (NM) and iron contrast ratio (CR) and normalized volume (nVol) for healthy controls (HCs) and PD patients according to the quantification threshold for the selection of hyper-intense or hypo-intense voxels in the case of NM and iron, respectively, in the automatically segmented nigrosome-1 (N1). Note that iron CR values have been inverted in sign to facilitate interpretation –higher iron CR means darker appearance in the image-, whereas NM CR values have not been modified –higher NM CR means brighter appearance in the image-. HC and PD groups were compared for each threshold by means of a Mann-Whitney's U test. Resulting p-values are represented in the right axis of *a-b* and *d-e*. Statistical significance value of p=.05 is also drawn as a horizontal line of reference. The optimal threshold for each parameter, corresponding to the minimum p-value between HC and PD, is also annotated in the figure. The number of HC or PD subjects with no NM or iron voxels detected in the N1 for each quantification threshold value was also calculated. **a)** N1 NM CR; **b)** N1 NM nVol; **c)** Number of control and PD subjects with no N1 NM voxels detected for each threshold; **d)** N1 iron CR; **e)** N1 iron nVol; **f)** Number of control and PD subjects with no N1 iron voxels detected for each threshold. Note that NM and iron nVol optimal quantification thresholds to maximize HC vs. PD differences were 0.24 and 0.30, respectively, but 0.22 and 0.20 values were selected to avoid having subjects with no NM or iron voxels detected in the N1.

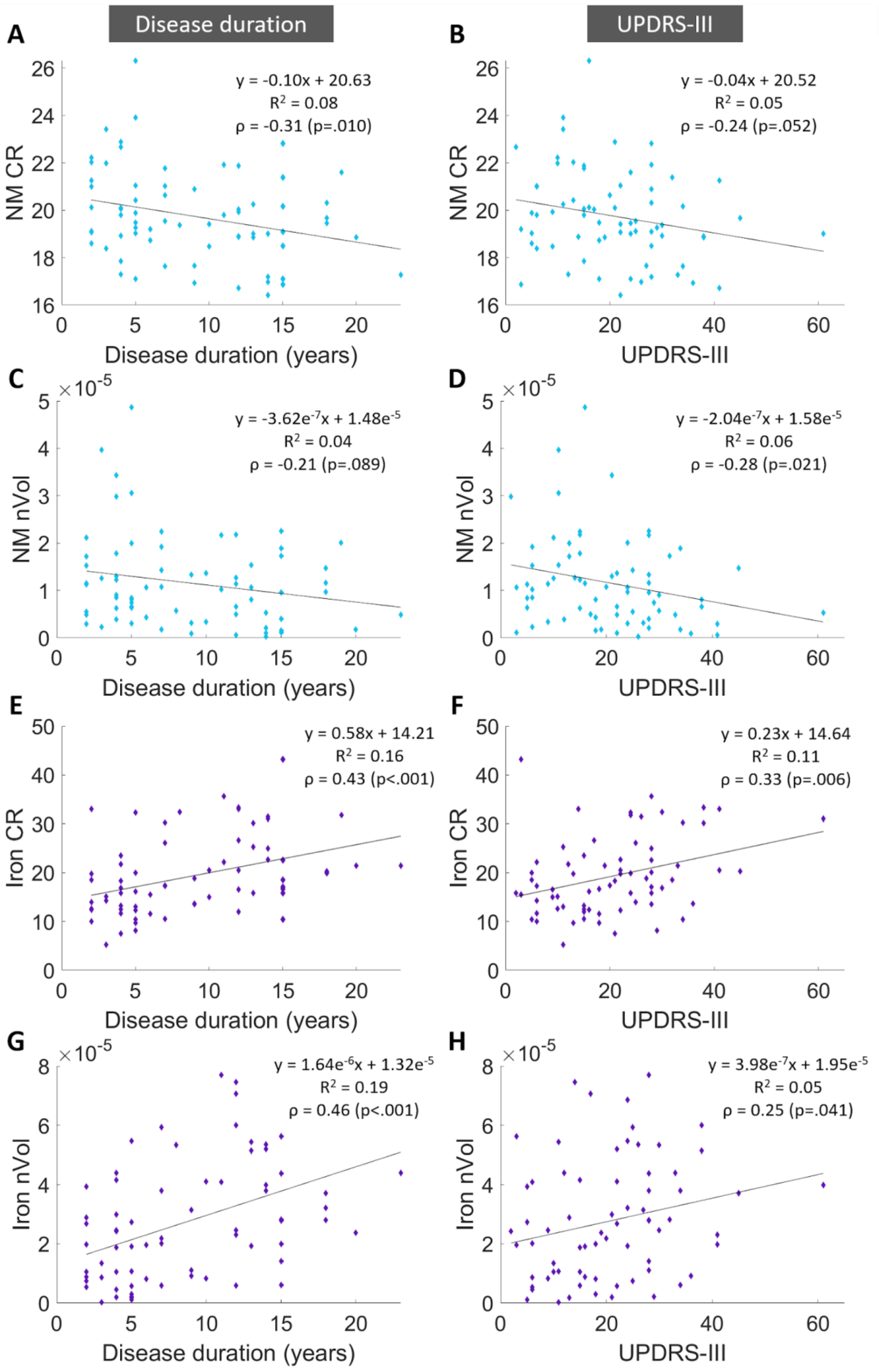
Disease duration
UPDRS-III
A
y = -0.10x + 20.63
$R^2$ = 0.08
ρ = -0.31 (p=.010)
NM CR
Disease duration (years)
B
y = -0.04x + 20.52
$R^2$ = 0.05
ρ = -0.24 (p=.052)
UPDRS-III
C
y = -3.62e-7x + 1.48e-5
$R^2$ = 0.04
ρ = -0.21 (p=.089)
NM nVol
D
y = -2.04e-7x + 1.58e-5
$R^2$ = 0.06
ρ = -0.28 (p=.021)
E
y = 0.58x + 14.21
$R^2$ = 0.16
ρ = 0.43 (p<.001)
Iron CR
F
y = 0.23x + 14.64
$R^2$ = 0.11
ρ = 0.33 (p=.006)
G
y = 1.64e-6x + 1.32e-5
$R^2$ = 0.19
ρ = 0.46 (p<.001)
Iron nVol
H
y = 3.98e-7x + 1.95e-5
$R^2$ = 0.05
ρ = 0.25 (p=.041)

**Supplementary Figure 2. Partial correlation analyses of nigrosome-1's image features with clinical variables of Parkinson's disease, using age and sex as covariates.** Correlation of N1's NM and iron CR and nVol of PD patients with disease duration and UPDRS-III. The Spearman's rank correlation coefficient (i.e., ρ) was calculated for each image-clinical parameter combination. Age and sex were used as covariates in the analysis, since they both affect the phenotypical expression of PD, and p-values were adjusted for comparisons through a multivariate permutation test. Corrected p-values <0.05 were considered statistically significant. **A)** Significant negative correlation between N1 NM CR and disease duration (p=.010). **B)** Non-significant negative correlation between N1 NM CR and UPDRS-III (p=.052). **C)** Non-significant negative correlation between N1 NM nVol and disease duration (p=.089). **D)** Significant negative correlation between N1 NM nVol and UPDRS-III (p=.021). **E)** Significant positive correlation between N1 iron CR and disease duration (p<.001). **F)** Significant positive correlation between N1 iron CR and UPDRS-III (p=.006). **G)** Significant positive correlation between N1 iron nVol and disease duration (p<.001). **H)** Significant positive correlation between N1 iron nVol and UPDRS-III (p=.041).

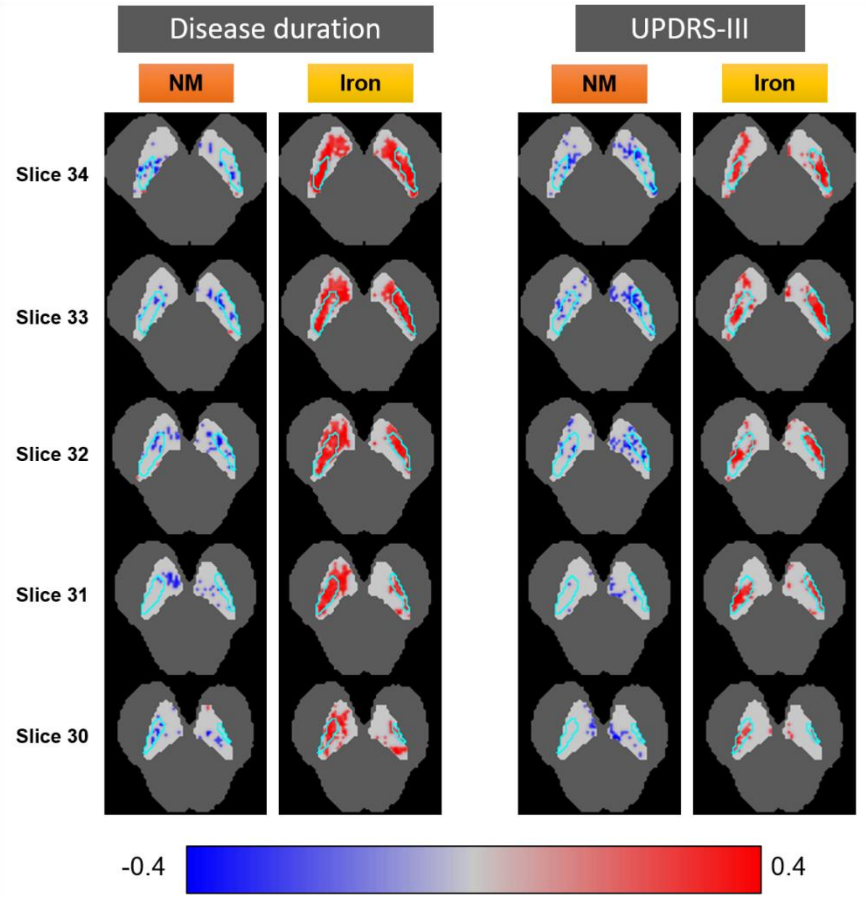


**Supplementary Figure 3. Statistically significant voxel-wise correlations between clinical scores and NM and iron in the N1 and SNc.** 2D axial slices of voxel-wise correlations between the CR of NM and iron of each SNc voxel, and disease duration (left) and UPDRS-III (right). The N1 region is outlined in cyan, and SNc and brainstem templates are shown in light grey and dark grey, respectively, as reference. Partial correlation analysis was performed using age and sex as covariates, and a multivariate permutation test was used to adjust the p-values to account for multiple voxels and multiple clinical scores. A blue/red colormap is used to display the value of the Spearman's rank partial correlation coefficient of each voxel that shows a significant correlation ($p<0.05$). Red indicates positive correlation between voxel signal and clinical score, whereas blue indicates negative correlation.

| Individual Parameter | AUC | Confidence Interval | p-value |
|---|---|---|---|
| N1 NM CR | 0.878 | [ 0.794 - 0.932 ] | <0.001 |
| N1 NM nVol | 0.863 | [ 0.774 - 0.930 ] | <0.001 |
| N1 iron CR | 0.878 | [ 0.797 - 0.933 ] | <0.001 |
| N1 iron nVol | 0.846 | [ 0.741 - 0.912 ] | <0.001 |
| SNc NM CR | 0.866 | [ 0.766 - 0.926 ] | <0.001 |
| SNc NM nVol | 0.910 | [ 0.838 - 0.956 ] | <0.001 |
| SNc iron CR | 0.814 | [ 0.710 - 0.886 ] | <0.001 |
| SNc iron nVol | 0.767 | [ 0.646 - 0.850 ] | <0.001 |
| **Regression Model** | **AUC** | **Confidence Interval** | **p-value** |
| *LogReg* [ N1 NM ] | 0.879 | [ 0.803 - 0.939 ] | <0.001 |
| *LogReg* [ N1 iron ] | 0.892 | [ 0.817 - 0.945 ] | <0.001 |
| *LogReg* [ N1 ] | 0.931 | [ 0.851 - 0.969 ] | <0.001 |
| *LogReg* [ SNc NM ] | 0.911 | [ 0.843 - 0.955 ] | <0.001 |
| *LogReg* [ SNc iron ] | 0.823 | [ 0.727 - 0.899 ] | <0.001 |
| *LogReg* [ SNc ] | 0.938 | [ 0.858 - 0.973 ] | <0.001 |
| *LogReg* [ N1 + SNc ] | 0.947 | [ 0.885 - 0.977 ] | <0.001 |

**Supplementary Table 1. Diagnostic performance results for HC vs. PD discrimination using image parameters automatically calculated for N1 and SNc.** Individual parameters are presented first, followed by the diagnostic performance of each brainstem structure obtained by combining their individual parameters (NM, iron, or both) through binary logistic regression. Finally, a complete model combining the parameters of both structures is assessed. Abbreviations: N1, nigrosome-1; NM, neuromelanin; CR, contrast ratio; nVol, normalized volume; SNc, substantia nigra pars compacta; *LogReg*, binary logistic regression.
